# Factors Associated with Newborn Hearing Screening Outcomes in a North Carolina Population

**DOI:** 10.1101/2025.08.02.25332199

**Authors:** Martin Moya, Maria E. Diaz-Gonzalez de Ferris, Steve Parsons

## Abstract

**Introduction:** This study aims to investigate factors that may impact newborn hearing screening (NHS) results.

**Methods:** A retrospective cohort study analyzed NHS results performed at UNC Southeastern Hospital between January 1, 2023, and June 30, 2024. Logistic regression analyses were performed to identify factors associated with NHS failure, including language spoken at home, race/ethnicity, sex, mode of delivery, birth weight, Apgar scores, and bilirubin levels. Stepwise selection methods were used to determine significant predictors.

**Results:** Among 2,642 neonates screened, 96.47% passed the initial hearing screening, while 3.53% failed and required follow-up testing. In univariate analysis, NHS failure was associated with vaginal delivery compared to C-section (4.14% vs. 2.11%, p = 0.0165). Male neonates had a higher failure rate compared to females (4.44% vs. 2.63%, p = 0.0205). Hispanic neonates vs. non-Hispanics had a greater failure rate (5.59% vs. 3%, p = 0.0046). Infants of Spanish-speaking mothers (6.1%) had a greater NHS failure (p = 0.016) compared to those of English-speaking mothers (3.07%) or Haitian-speaking mothers (4.08%). In multivariate analysis, stepwise backward elimination identified ethnicity (p = 0.0054) and bilirubin levels (p = 0.049) as independent predictors of NHS failure, with a joint model significance of p = 0.027.

**Conclusions:** This study suggests that mode of delivery, maternal language, ethnicity, and bilirubin levels may influence NHS outcomes. Contrary to some previous reports, vaginally delivered infants had a higher failure rate than those born via C-section. Understanding these influences may optimize screening protocols, reduce unnecessary referrals, and mitigate parental anxiety.

## INTRODUCTION

Hearing loss in early life has a significant health concern, impacting a child’s development in language, cognition, and social-emotional well-being(1). The linguistic and communicative abilities of children with hearing loss can be maximized by early detection and rehabilitation to ensure their successful integration into a hearing society(2). Early detection of hearing abnormalities is crucial for timely intervention, and newborn hearing screening programs facilitate timely intervention.

In our own Neonatal Intensive Care Unit (NICU), anecdotal observations by our nursing staff have indicated a potential trend of increased NHS failures among infants born to Spanish-speaking mothers. This study aims to investigate the potential influence of maternal language spoken at home, mode of delivery, race/ethnicity, and other perinatal factors on risk factors for abnormal NHS results in patients served at the poorest county in North Carolina.

## METHODS

### Study Design, Setting and Ethics

This retrospective cohort study was performed on all consecutive neonates delivered at the University of North Carolina Southeastern Regional Hospital between January 1, 2023, and June 30, 2024. This 18-month study was approved by The University of North Carolina, Office of Human Research Ethics, which waived obtaining written consent from the parents of the neonates given the nature of the data analysis.

### Study Population

This study included 2642 AABR tests in all infants delivered at our institution during the study period regardless of their gestational age or postnatal unit (well-baby or neonatal intensive care -NICU-). No infants were excluded due to incomplete or corrupted data, and no infants were presented with craniofacial anomalies that would preclude accurate hearing screening.

### Neonatal Hearing Screening

Automated Auditory Brainstem Response (AABR) tests (GSI AUDIOscreener+; Grason-Stadler, Minneapolis, MN) were used for the hearing screening prior to discharge from the hospital. Infants requiring admission to the Neonatal Intensive Care Unit (NICU), including those born preterm or with other medical conditions necessitating NICU care, were screened with AABR prior to discharge from the NICU. Infants who did not pass the initial AABR in either ear or both ears were scheduled for a retest approximately two weeks following the initial screening.

The response detection method used in our automated AABR device is termed Fsp and is designed to estimate the ratio of a signal (AABR) to background noise for a given averaged AABR(3). The newborns were naturally asleep, and no medication was used when the test was conducted. We used 35 dB HL click sound at a rate of 37.1/s for the stimulus. Point-optimized variance ratios higher or less than 3.5, were designated as “pass” or “refer,” respectively. This intensity level was used in infants as well as in NICU infants to ensure that the neural response could be recorded, unaffected by any possible conductive hearing loss caused by postnatal debris in the external ear canal or by middle ear fluids.

### Data Collection

Data were obtained from Early Hearing Detection and Intervention Program (EHDI), Division of Child and Family Well-Being, North Carolina Department of Health and Human Services, as well as from the UNC Southeastern Hospital newborn hearing screening database. The data collected included infant demographics (gestational age, birth weight, height, head circumference, sex, race/ethnicity), maternal demographics (age, language spoken at home, mode of delivery, medications, administered during labor), AABR results (pass/refer for each ear), and date and time of screening.

Data integrity and accuracy were verified by the nurse in charge of the hearing screening program at our hospital. This highly experienced and trained nurse is responsible for conducting the screenings, ensuring their quality, guaranteeing data accuracy and proper data entry, and training other nurses who work alongside her in the program

### Statistics

Data analysis was performed using [JMP Pro 17 (SAS Institute Inc., Cary, NC, USA)]. Continuous variables are presented as mean ± standard deviation (SD) or median [interquartile range (IQR)] depending on their distribution, which was assessed using Shapiro-Wilk test. Categorical variables are presented as frequencies and percentages.

To determine the factors associated with failing the hearing screening (HS), multiple logistic regression analyses were conducted. Initially, a comprehensive analysis was performed with “pass/fail” on the NHS as the dependent variable and 19 independent variables, including language, race, ethnicity, sex, mode of delivery, birth weight, length, head circumference, blood type, pH, PCO2, HCO3, Apgar at 1, 5 and 10 minutes, hemoglobin, bilirubin, neutrophil count and glucose. A standard least squares analysis with minimal reporting was initially conducted. Following the parameter estimates, the variance inflation factor (VIF) was calculated to identify unstable variables and those exhibiting multicollinearity. A VIF threshold of 40 was used to remove variables. With the remaining variables, a correlation analysis was performed on the numerical variables to assess multicollinearity. No correlation greater than 80% was found between any of these numerical variables, and therefore all were retained in the model. Subsequently, the variables were analyzed using a stepwise forward selection method with varying entry thresholds (0.05, 0.10, and 0.20). A stepwise backward elimination analysis was also performed using a threshold of 0.10.

## RESULTS

We analyzed data from a cohort of 2,642 neonates representing a wide range of socio-demographic and clinical profiles. Table 1 presents the distribution of key variables among neonates who passed the hearing screening compared to those who did not.

**Table 1.**

| Variables | Pass the Hearing screening | Do Not pass the hearing screening | Statistics |
| --- | --- | --- | --- |
| Weight | 3232.4 ± 11 | 3282.1 ± 57.7 | 0.4 |
| Height | 51.5 ± 0.1 | 52.3 ± 0.7 | 0.23 |
| Head Circumference | 33.6 ± 0.1 | 33.5 ± 0.4 | 0.84 |
| Blood Type |  |  |  |
| O Positive | 68.2% | 82.3% | 0.52 |
| A Positive | 15.7% | 9.7% |  |
| B Positive | 7.6% | 3.2% |  |
| AB Positive | 0,8% | 0 |  |
| O Negative | 4,8% | 3.2% |  |
| A Negative | 1.7% | 1.6% |  |
| B Negative | 1% | 0 |  |
| AB Negative | 0.2% | 0 |  |
| Abs Neutr Value | 6575 ± 254 | 5111 ± 1561 | 0.35 |
| Hemoglobin | 14.7 ± 0.2 | 14.2 ± 1.2 | 0.63 |
| Bilirubin | 6.3 ± 0.1 | 7.3 ± 0.5 | 0.16 |
| Glucose | 83.1 ± 1.7 | 90.4 ± 9.9 | 0.46 |
| Blood gases |  |  |  |
| pH | 7.35 ± 0.01 | 7.43 ± 0.08 | 0.38 |
| CO2 | 43.2 ± 2.7 | 37.7 ± 17.6 | 0.76 |
| HCO3 | 22.6 ± 0.4 | 24.9 ± 2.5 | 0.34 |
| Apgar |  |  |  |
| 1 minute | 7.8 ± 0.03 | 7.9 ± 0.13 | 0.89 |
| 5 minutes | 8.8 ± 0.01 | 8.8 ± 0.07 | 0.89 |
| 10 minutes | 8.6 ± 0.1 | 8.5 ± 0.48 | 0.9 |

With respect to biometric parameters, no statistically significant differences were observed between groups in birth weight, length, or head circumference. Similarly, hematologic indices, including absolute neutrophil count, hemoglobin concentration, bilirubin levels, and glucose levels, were comparable across both groups.

Analysis of arterial blood gas parameters revealed no significant intergroup differences in pH (p = 0.38), partial pressure of carbon dioxide (CO_2_; p = 0.76), or bicarbonate (HCO_3_^−^; p = 0.34). Apgar scores assessed at 1, 5, and 10 minutes post-delivery did not differ significantly between groups (p > 0.05). Additional clinical variables evaluated included gestational age, type of care unit (neonatal intensive care unit [NICU] vs. well-baby unit), timing of auditory brainstem response (ABR) testing, diagnosis of hypoxic-ischemic encephalopathy (HIE), and exposure to perinatal medications. None of these variables demonstrated statistically significant differences between neonates who passed and those who did not pass the hearing screening.

Analysis of demographic and perinatal characteristics revealed statistically significant associations with newborn hearing screening outcomes, as illustrated in **Figure 1**. Ethnicity emerged as a significant factor influencing screening results (**p = 0.0046**). A notably higher proportion of neonates identified as Hispanic did not pass the initial hearing screening (35.7%) compared to those who passed (22.5%), suggesting a potential disparity in early auditory outcomes among ethnic groups (Figure 1A).

**Figure 1.**
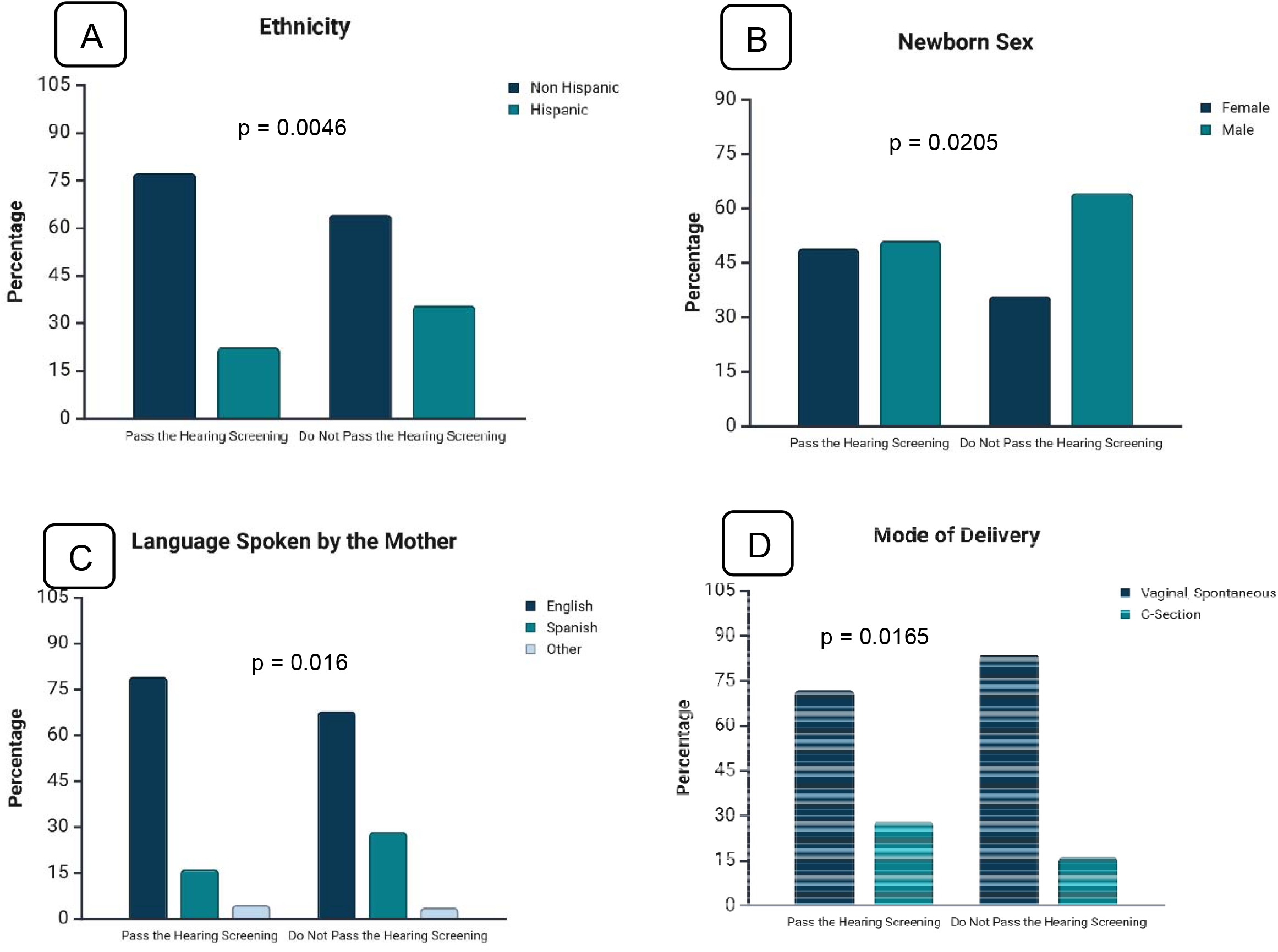
This figure presents newborn hearing screening outcomes in relation to various maternal and birth-related factors. Panel A illustrates pass rates by infant ethnicity (Non-Hispanic vs. Hispanic). Panel B shows pass rates by newborn sex (Female vs. Male). Panel C depicts pass rates based on the primary language spoken by the mother (English vs. Spanish vs. Other). Panel D presents pass rates categorized by mode of delivery (Vaginal Spontaneous Delivery vs. C-Section).

Sex-based differences were also observed, with a statistically significant variation in screening outcomes between male and female neonates (**p = 0.0205**). Male infants constituted a greater proportion of the non-passing group (64.2%) relative to the passing group (51.1%), indicating a possible sex-related vulnerability in early auditory function or screening sensitivity (Figure 1B).

Maternal primary language demonstrated a significant association with screening performance (**p = 0.016**). While English was the predominant language spoken by mothers in both outcome groups, neonates born to Spanish-speaking mothers exhibited a higher rate of screening failure (28.4%) compared to those whose mothers primarily spoke English (16.2%). This finding may reflect underlying sociolinguistic or healthcare access factors that influence early hearing outcomes (Figure 1C).

Furthermore, mode of delivery was significantly correlated with screening results (**p = 0.0165**). A higher proportion of neonates delivered via spontaneous vaginal delivery (SVD) failed the hearing screening (83.7%) compared to those who passed (71.9%). This association may not be attributable to transient middle ear conditions, such as fluid retention, less commonly observed following vaginal birth, which can affect otoacoustic emission or automated auditory brainstem response testing (Figure 1D).

Collectively, these findings underscore the influence of ethnicity, neonatal sex, maternal language, and delivery mode on the outcomes of universal newborn hearing screening.

To explore which clinical and demographic variables were most strongly associated with failure in the newborn hearing screening, a series of logistic regression analyses were conducted. An initial model incorporating 19 variables—including neonatal anthropometrics, maternal language, ethnicity, delivery mode, and various laboratory and clinical parameters—was refined through multicollinearity assessment using variance inflation factors (VIF), leading to the exclusion of 8 variables. Among the remaining 11, stepwise forward selection identified race and mode of delivery as potential predictors at a 0.10 significance threshold, though only mode of delivery remained statistically significant in the final model (p = 0.0153).

To complement this, a stepwise backward elimination approach was applied using the same entry threshold. This method initially retained six variables—race, bilirubin, neutrophil count, glucose, delivery mode, and ethnicity. After fitting the model to the dataset, only race (p = 0.0054) and bilirubin (p = 0.049) remained statistically significant, with a joint model p-value of 0.027. These results suggest that, in addition to delivery mode, both racial background and neonatal bilirubin levels may independently contribute to the likelihood of a newborn not passing the initial hearing screening.

## DISCUSSION

This study aimed to identify clinical and sociodemographic factors associated with failure in newborn hearing screening (NHS) among neonates born at UNC Southeastern Hospital between January 2023 and June 2024. The investigation was prompted by observations from nursing staff who noted a potential trend of increased NHS failures among infants born to Spanish-speaking mothers. In a cohort of 2,642 infants screened using automated auditory brainstem response (AABR), 3.53% did not pass the initial screening. Univariate analyses revealed that vaginal delivery, male sex, Hispanic ethnicity, and maternal Spanish language were significantly associated with higher NHS failure rates. Multivariate logistic regression using stepwise selection methods further identified mode of delivery, race, and bilirubin levels as independent predictors of screening failure. These findings suggest that both perinatal and sociodemographic variables may influence NHS outcomes and highlight the need for tailored screening strategies to reduce disparities and improve early detection of hearing loss.Newborn hearing screening programs are crucial for early identification of hearing loss, enabling timely intervention and improving language development outcomes. While NHS programs have become standard practice, various factors can influence screening outcomes, leading to false positives, increased healthcare costs, and heightened parental concern. This discussion explores the potential impact of delivery mode and sociodemographic factors on NHS results, highlighting areas of ongoing investigation and the need for further research.

Several studies have explored the association between delivery methods and NHS outcomes. Some research suggests that infants born via cesarean section (C-section) exhibit higher rates of “refer” or “fail” results on initial hearing screenings compared to vaginally delivered infants(4) (5) (6) (7). The mechanisms underlying this association are not fully understood, but potential explanations include retained amniotic fluid in the middle ear of C-section infants, which can interfere with sound conduction(8), and transient physiological changes related to the birthing process itself. Our findings, illustrated in Figure 1D, do not support this observation, showing a higher “not pass” rate for infants delivered vaginally (83.7%) compared to the “pass” rate of 71.9% for the same mode of delivery(9). Additionally, the influence of anesthesia warrants consideration. A recent study by Guler et al. (2025)(10) specifically investigated the impact of both delivery type and anesthesia method on Auditory Brainstem Response test results, finding a statistically significant relationship between these factors and initial screening outcomes, particularly in newborns delivered by C-section under general anesthesia.

Conversely, it is important to acknowledge that other studies have found no significant association between delivery mode and NHS results(11). Furthermore, research has also suggested that certain medications administered during labor, such as pethidine, may contribute to false-positive results in newborn hearing screenings(12). To address this potential confounder, we specifically examined the use of pethidine within our obstetric service. Our review confirmed that this medication has not been administered in our facility for several years. This finding suggests that pethidine exposure is unlikely to have influenced our results, reinforcing the need to explore alternative explanations for the observed disparities.

Our analysis also revealed disparities in NHS outcomes based on sociodemographic factors. Among infants who passed the hearing screening, 16.2% had mothers who primarily spoke Spanish. In contrast, this proportion increased to 28.4% among infants who did not pass, a statistically significant difference (p = 0.016), as shown in Figure 1C. Similarly, Figure 1A shows that among infants who passed the hearing screening, 22.5% were Hispanic. In contrast, this proportion increased to 35.7% among infants who did not pass, indicating a trend toward higher failure rates among Hispanic infants. This observation, while preliminary, raises important questions about the potential influence of factors such as race, ethnicity, and primary language spoken in the home on NHS outcomes. These factors may interact with other perinatal variables and contribute to disparities in screening results. Further investigation is needed to understand the underlying reasons for these disparities. Potential contributing factors could include differences in access to prenatal care and health education, cultural beliefs and practices surrounding healthcare and early childhood development, and socioeconomic disparities that may affect access to resources and quality of care.

The discrepancies in findings across existing studies, coupled with our observations, highlight the need for further research to clarify the complex interplay between delivery methods, sociodemographic factors, and NHS outcomes. Understanding these influences is essential for optimizing screening protocols, minimizing unnecessary referrals, and reducing parental stress, particularly within diverse populations. Future research should focus on larger, more diverse samples to draw more robust conclusions, as well as longitudinal studies to track the impact of early screening results on long-term language and developmental outcomes. Additionally, investigating the specific mechanisms by which delivery mode and sociodemographic factors influence NHS outcomes is crucial. By addressing these research gaps, we strive to improve the accuracy and effectiveness of NHS programs, ensuring that all newborns have the opportunity for early identification and intervention for hearing loss.

It is important to consider that differences in delivery methods between Latina and Non-Latina mothers could act as a potential confounder in our findings. To account for this, we employed propensity score matching (PSM) to create a balanced subset in which Latina and Non-Latina mothers had comparable distributions of vaginal and cesarean deliveries. This method reduces confounding by simulating a randomized controlled trial-like scenario, ensuring that delivery mode does not bias our results. After matching, we reanalyzed hearing screening outcomes and found that the observed disparities between the groups remained statistically significant. This strengthens our findings and suggests that factors beyond the mode of delivery contribute to the observed differences.

The discrepancies in findings across existing studies, coupled with our observations, highlight the need for further research to clarify the complex interplay between delivery methods, sociodemographic factors, and NHS outcomes. Understanding these influences is essential for optimizing screening protocols, minimizing unnecessary referrals, and reducing parental stress, particularly within diverse populations. Future research should focus on larger, more diverse samples to draw more robust conclusions, as well as longitudinal studies to track the impact of early screening results on long-term language and developmental outcomes. Additionally, investigating the specific mechanisms by which delivery mode and sociodemographic factors influence NHS outcomes is crucial. To minimize potential biases associated with our specific patient population, we have requested access to statewide statistics from the North Carolina Department of Health and Human Services. By analyzing this broader dataset, we aim to validate our findings and gain a more comprehensive understanding of the factors affecting newborn hearing screening outcomes in North Carolina.

## Data Availability

All data produced in the present study are available upon reasonable request to the authors

